# Is e-cigarette use in non-smoking young adults associated with later smoking? A systematic review and meta-analysis

**DOI:** 10.1101/19007005

**Authors:** Jasmine N Khouja, Steph F Suddell, Sarah Peters, Amy E Taylor, Marcus R Munafò

**Author notes:** Correspondence to: Jasmine N. Khouja; 12a Priory, Road, University of Bristol, Bristol, UK, BS8 1TU.

## Abstract

**Objective:** The aim of this review was to investigate whether e-cigarette use compared to non-use in young non-smokers is associated with subsequent cigarette smoking.

**Data sources:** PubMed, Embase, Web of Science, Wiley Cochrane Library databases, and the 2018 Society for Research on Nicotine and Tobacco and Society for Behavioural Medicine conference abstracts.

**Study selection:** All studies of young people (up to age 30 years) with a measure of e-cigarette use prior to smoking and an outcome measure of smoking where an odds ratio could be calculated were included (excluding reviews and animal studies).

**Data Extraction:** Independent extraction was completed by multiple authors using a pre-prepared extraction form.

**Data synthesis:** Of 9,199 results, 17 studies were included in the meta-analysis. There was strong evidence for an association between e-cigarette use among non-smokers and later smoking (OR 4.59, 95% CI 3.60 to 5.85) when the results were meta-analysed in a random effects model. However, there was high heterogeneity (*I*^*2*^ = 88%).

**Conclusions:** Whilst the association between e-cigarette use among non-smokers and subsequent smoking appears strong, the available evidence is limited by the reliance on self-report measures of smoking history without biochemical verification. None of the studies included negative controls which would provide stronger evidence for whether the association may be causal. Much of the evidence also failed to consider the nicotine content of e-liquids used by non-smokers meaning it is difficult to make conclusions about whether nicotine is the mechanism driving this association.

## Introduction

Existing evidence suggests that electronic cigarette (e-cigarette) use is less harmful than smoking ^1^ and is an effective smoking cessation aid ^2^; however, there are concerns that e-cigarettes may act as a gateway to smoking cigarettes among young people. If this is correct, rather than seeing a decline in smoking rates we may see smoking rates remaining stable or increasing due to a new generation of smokers for whom e-cigarettes have acted as a route into smoking. This hypothesis (sometimes referred to as the ‘gateway hypothesis’ or ‘catalyst model’) has been widely debated among researchers and public health officials. Some argue that a common liability better explains the association between vaping and later smoking, whereby the same genetic or environmental factors that increase the likelihood of someone vaping also increase the likelihood of someone smoking.^3^ As many people use e-cigarettes to help them stop smoking, it would also be logical to assume the opposite direction of causality with smoking causing people to vape.^4^ The lack of consensus on the issue demonstrates the need for the current evidence to be synthesised.

One systematic review and meta-analysis of the association between baseline e-cigarette use and later smoking concluded that e-cigarette use was associated with an increased likelihood of smoking at follow up.^5^ Although this meta-analysis is relatively recent, this is a fast-moving field with a substantial number of relevant studies having been published since 2017. Given the topic is of great interest to researchers and policy makers, an updated meta-analysis is necessary. Moreover, in the previous meta-analysis,^5^ moderate heterogeneity was observed between the study results. Some potential sources of heterogeneity could include the age range of the participants in the studies and risk of bias among the studies. Soneji and colleagues ^5^ addressed this by stratifying by average age, finding that there was greater heterogeneity between studies of adolescents (under 18 years) compared to studies of young adults. However, they did not stratify by risk of bias. This is important because the preconceptions of study authors may also influence how studies are designed and conducted, and this may be reflected in a study’s conclusions. For example, two studies in the Soneji and colleagues ^5^ meta-analysis drew diverging conclusions, despite the pooled odd ratios not differing substantially from each other. Leventhal and colleagues ^6^ concluded there was insufficient evidence to support the gateway hypothesis, whereas Miech and colleagues ^7^ concluded that there was a one-way bridge from e-cigarette use to smoking, despite both studies having similar results.

In this systematic review and meta-analysis, we updated and extended previous reviews in which e-cigarette use and later smoking have been explored while looking at a broad range of evidence. Our aim was to investigate whether e-cigarette use, compared to non-use, in young non-smokers is associated with subsequent cigarette use by combining evidence from studies investigating e-cigarette use and subsequent smoking where an odds ratio could be calculated. Additionally, we aimed to use stratification to explore sources of heterogeneity and biases in conclusions regarding the gateway hypothesis. From our knowledge of the evidence base and considering this is a fast-moving area of research, we expected to identify a substantial number of studies that have been published since the review by Soneji and colleagues.^5^

## Methods

The protocol for this systematic review and meta-analysis was published online prior to initiating the search and can be found on the Open Science Framework (https://osf.io/3gc2y/). PRISMA and MOOSE guidelines were followed (where applicable).

### Eligibility Criteria

We included randomised controlled trials, longitudinal studies, cross-sectional studies, and case-control studies. Only studies investigating young people aged up to the age of 30 years old (inclusive) were included. Studies had to have a baseline or retrospective measure of e-cigarette use (including but not limited to ever, occasional, heavy, recent, regular or frequent use) prior to initiating smoking and a measure of cigarette smoking (including but not limited to ever, occasional, heavy, recent, regular, frequent or escalated smoking) as an outcome. Studies had to include a comparison group (i.e., group which the exposed group is compared to), which could include young people who were never, trial or not recent e-cigarette users or smokers, dependent on the study. Review articles and animal studies were excluded.

### Information Sources

Our search strategy was a replication and extension of a strategy used in a similar review.^5^ We conducted an electronic search of the databases PubMed, Embase, Web of Science, Wiley Cochrane Library, Society for Research on Nicotine and Tobacco, and the Society for Behavioural Medicine. Due to member restricted access we were unable to search the NIH Tobacco Regulatory Science Conference abstracts as stated in our pre-registered protocol. We compared the list of studies to be included to those included in previous similar reviews to ensure the search strategy had not omitted any relevant studies. Studies written in languages other than English were translated by colleagues and using Google translate where translations were not already available.

The search strategy was conducted up to 24^th^ November 2018. E-cigarettes are a relatively new product on the consumer market; therefore, no date restrictions were placed on the search strategy.

### Search Strategy

Studies were initially selected for screening using the following search terms within the titles, abstracts or keywords: Cigar* OR Tobacco OR Smok* AND Electronic Cigarette* OR E-Cig* OR Electronic Nicotine Delivery System* OR Vape OR Vaping OR Alternative Nicotine Delivery System*. Boolean operators and truncations differed depending on the database. Relevant MeSH terms were included when searching the PubMed database. An example search can be found in the Supplementary Material.

### Study Selection and Data Collection Process

Study selection and data extraction took place over three stages. Stage 1 consisted of title and abstract screening; Stage 2 consisted of a full text screening; Stage 3 consisted of data extraction from selected studies. A full list of extracted data is included in the supplementary material. Titles, abstracts and full text articles were double screened and then double extracted by three reviewers (JK, [100%], SS [50%], and SP [50%]). Discrepancies were resolved by a third reviewer where necessary.

Covidence (www.covidence.org), an online systematic review tool which is in partnership with Cochrane, was used to streamline and document this process.

When insufficient information was available to determine eligibility, we contacted study authors. Where insufficient information was provided or obtained, the text was excluded from the review.

### Risk of Bias Assessment

Risk of bias was assessed using the Newcastle-Ottawa Scale (NOS). The selection, comparability and outcome domains of the tool were used to assess the risk of bias in all full texts included in the review. The studies were rated as good, fair or poor quality based on the star system of the tool (maximum of 9 stars, see supplementary material for more information). Quality/risk was double assessed by the review team. Studies were not excluded based on risk of bias.

Risk of bias across studies was assessed using the symmetry and 95% confidence region of a funnel plot.^8^ Asymmetry and more than 5% of points lying above the 95% confidence region may indicate some bias across studies.

### Causality Assessment

Using Bradford-Hill criteria, we selected four criteria which were relevant to the studies of interest to indicate the strength of evidence of a possible causal relationship between our exposure and outcome (see Supplementary material for more details).

These were: strength of association, specificity, temporality and dose responsivity. These criteria are particularly relevant to studies assessing whether e-cigarettes may act as a gateway to smoking.^3^

### Summary Measures

Effect estimates were reported as odds ratios (and converted where necessary). Odds ratios of the association between e-cigarette use and later cigarette use were combined using a random effects model. All unadjusted odds ratios were calculated using observed data points which were obtained from the original study or directly from the author if insufficient information was provided in the original study. Calculated effect sizes were double checked by the review team.

Adjusted odds ratios were reported as they were in the original study and adjusted risk ratios were converted to odds ratios using a modified version of a formula published in the Cochrane Handbook (Section 12.5.4.4)^8^: OR = (-RR + RR × ACR) / (RR × ACR - 1), where OR = odds ratio; RR = risk ratio; ACR = assumed control risk (calculated on a per study basis as the risk of later smoking among controls).

### Synthesis and Results

In a random-effects model, we calculated the pooled odds ratios from unadjusted and adjusted odds ratios for ever cigarette use at follow up among never smokers at baseline, in ever compared to never e-cigarette users at baseline. Where multiple exposure or outcome measures were included in the original study, this estimate was used in the main analysis. If ever use of e-cigarettes was not reported, the main effect reported in the study was included in our main analyses.

Where possible (i.e., where more than one study was available), we also analysed the results in a series of subgroups – we pooled odds for: i) ever versus never e-cigarette use at baseline and ever versus never smoking at follow up, ii) ever versus never e-cigarette use at baseline and current (past 30 day) versus non-current use of cigarettes at follow up, and iii) current vs non-current e-cigarette use at baseline and ever vs never smoking at follow up. In retrospective studies, measures of e-cigarette use prior to smoking were treated as baseline and smoking status at the time of the study was treated as the follow up. We aimed to pool the results of regular (at least monthly for more than 6 months) cigarette use at follow up among never smokers at baseline, in regular compared to non-regular e-cigarette users at baseline; however, insufficient data were available to do so.

Heterogeneity of study effect estimates can be indicated by an *I*^*2*^ statistic. Sources of heterogeneity were explored through subgroup analysis. All analyses were conducted using Stata SE version 15.1 and Review Manager version 5.

### Patient and public involvement

This research was done without public involvement. The public was not invited to partake in the conception or design of the study, or the interpretation of the results. The public was not invited to contribute to the writing or editing of this document for readability or accuracy. The results of this meta-analysis will be disseminated to the public as widely as possible.

## Results

### Study Selection

Figure 1 shows the PRISMA study selection flow chart. A total of 15,519 studies were selected for title and abstract screening, 9,199 remained after exclusion of duplicates. After title and abstract screening, 133 studies were selected for full text screening. Of these, 24 studies were initially selected for inclusion; however, 7 studies were not included in the meta-analysis because the data overlapped with other included studies. Where data overlapped, the most relevant study was selected based on aims (i.e., studies where the primary aim addressed the question of interest were selected above those which addressed the question in secondary analysis) and sample size (i.e., larger sample sizes were included where both studies were relevant). In the meta-analysis, 16 studies were included in the main pooled unadjusted analysis and 17 studies were included in the pooled adjusted analysis. One study was excluded from the unadjusted analysis due to insufficient raw data availability but had adjusted results available.

**Figure 1.**
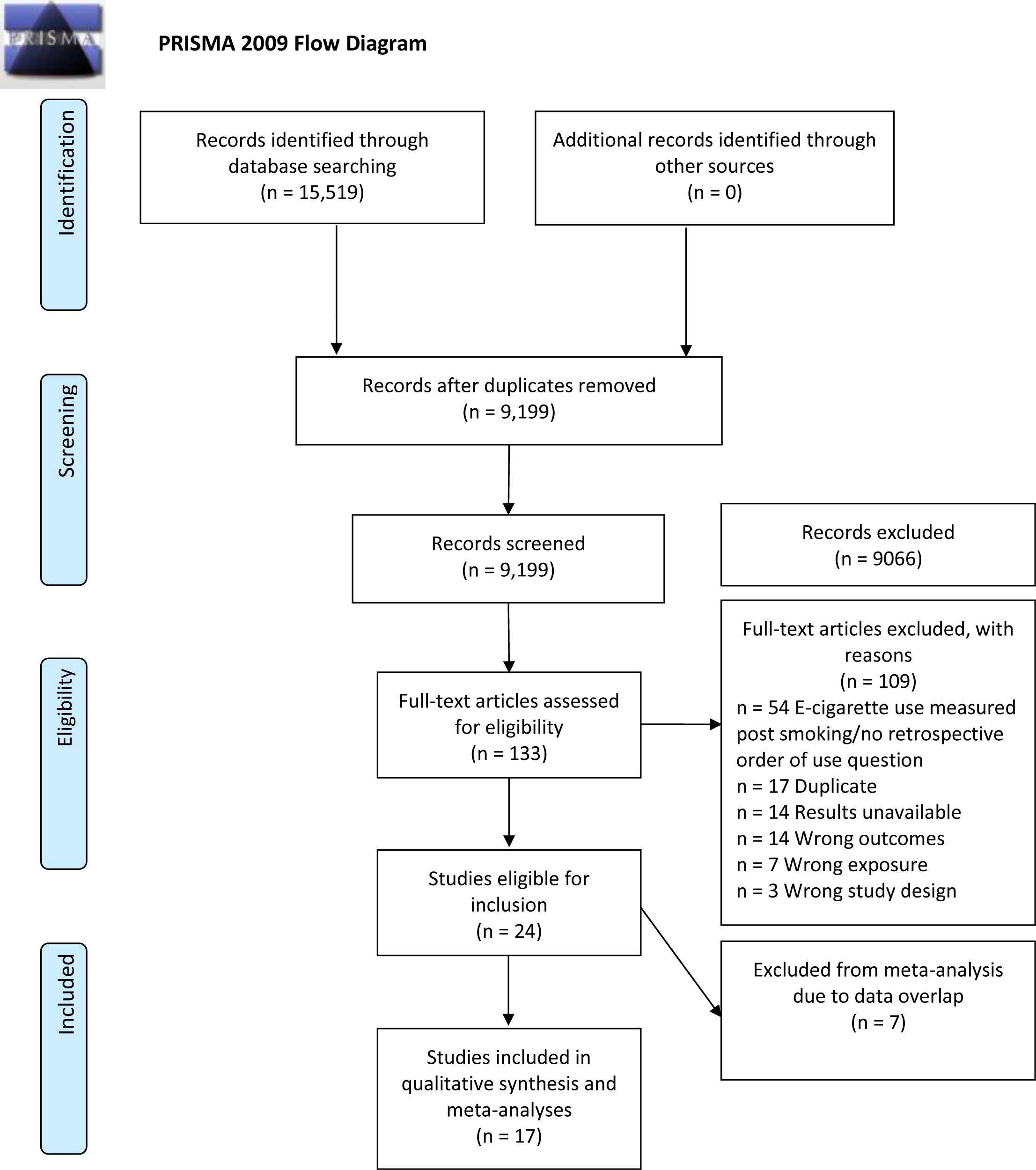
Study selection flow diagram.

### Study Characteristics

Details of the study characteristics are shown in Table S1. The majority of the studies included were longitudinal and one was cross-sectional in which participants were asked questions regarding their product use retrospectively. Total study sizes varied considerably, ranging from 347 to 39,718 and the number of participants included in the final analyses were often substantially smaller. Participants were mostly under 18 and many of the studies were school-based. Where the number of males and females in the study were reported, around 50% of participants were male in most cases. However, only 33% were male in one study.^9^ The majority of the studies (ten) were conducted in the US, three studies took place in the UK, one was based in Canada, one in Mexico, one in Germany and one in the Netherlands. Follow up periods ranged from 4-24 months.

In terms of observed exposures, most studies explored ever e-cigarette use with never e-cigarette users as a comparator. Two studies looked at current users with not current users as the comparator ^7 10^ and two looked at both current and ever use.^11 12^ Only one study considered the amount of exposure to nicotine,^13^ and one study looked at frequency of e-cigarette use.^14^ In the one study that took nicotine use into account,^13^ two separate analyses were conducted for: 1) ever use of nicotine containing e-cigarettes (OR = 11.90, 95% CI 3.36 to 42.11); and 2) ever use of non-nicotine containing e-cigarettes (OR= 5.36, 95% CI 2.73 to 10.52). However, the analysis groups were not mutually exclusive (i.e., an individual would have been in both analysis groups if they had tried both nicotine containing and nicotine free e-cigarettes). No analysis was reported using subgroups of exclusive nicotine or nicotine-free use.

The one study which addressed frequency of e-cigarette use ^14^ found that those who had used e-cigarettes at varying frequencies from once or twice (OR = 2.88, 95% CI 1.96 to 4.22) to weekly/daily (OR = 4.09, 95% CI 2.43 to 6.88) were more likely than those who had not used e-cigarettes to have smoked at least once at follow up.

Most of the included studies used ever smoking as an outcome. One study explored experimentation with smoking, as well as frequent and infrequent smoking,^15^ and three looked at recent/current smoking at follow up.^6 11 12^

Covariates included in the analyses varied greatly between studies. One study only adjusted for four covariates ^16^ while another adjusted for over 20 covariates.^17^ All studies adjusted for sex and most adjusted for age and race/ethnicity. Other frequently included covariates were peer smoking, sensation seeking (and related factors), and drug and alcohol use. No studies adjusted for nicotine exposure via e-cigarettes (i.e., e-liquid content and/or frequency of nicotine exposure).

### Quality/Risk of Bias Within Studies and Causality

The quality of studies (or inversely, risk of bias) was good in most cases when rated using the NOS (Table S2). One study was rated as fair quality,^6^ and three were rated as poor quality.^13 18 19^ Of the four Bradford-Hill criteria for causality deemed relevant to this research, the majority (11 studies) met three criteria (usually strength of evidence, temporality and specificity), four studies only met two criteria ^6 9 10 18^ and two met four criteria.^13 14^

## Results of Individual Studies

The results of individual studies included in the main meta-analysis can be found in Table 1 and within forest plots in figures 2 (unadjusted) and 3 (adjusted). Effect sizes (odds ratios [OR]) ranged from 2.39 to 12.31 (unadjusted). All estimates were considered to show strong evidence of a positive association between e-cigarette use among non-smokers and later smoking in unadjusted analyses. Covariates included in the adjusted analyses varied on a study-by-study basis. After adjustment, effects in all but two studies ^6 9^ remained strong. The inclusion of covariates in the model attenuated most results (although none were attenuated to the null). Effect sizes were strengthened after adjustment in four studies.^13 15 19 20^

**Table 1.** Individual results of studies included in the meta-analysis

| Study | Initiated cigarette smoking (n) |  | Odds of initiating smoking |  |
| --- | --- | --- | --- | --- |
|  | E-cigarette users | Never/not current e-cigarette users | Unadjusted odds ratio (95% CI) | Adjusted odds ratio (95% CI) |
| Auf, et al. <sup>18</sup> | * | * | * | 3.7 (3.1, 4.5) |
| Barrington-Trimis, et al. <sup>15</sup> | 184/857 | 280/4,171 | 3.80 (3.10, 4.66) | 4.57 (3.56, 5.87) |
| Best, et al. <sup>16</sup> | 74/183 | 249/1,942 | 4.62 (3.34, 6.38) | 2.42 (1.63, 3.60) |
| Conner, et al. <sup>29</sup> | 118/343 | 124/1,383 | 5.32 (3.99, 7.11) | 4.06 (2.94, 5.60) |
| East, et al. <sup>37</sup> | 11/21 | 74/902 | 12.31 (5.06, 29.94) | 10.57 (3.33, 33.50) |
| Hammond, et al. <sup>10</sup> | 136/487 | 1,313/16,831 | 4.58 (3.73, 5.63) | 2.12 (1.68, 2.66) |
| Leventhal, et al. <sup>6</sup> | 19/222 | 71/2,308 | 2.95 (1.74, 4.99) | 1.75 (1.10, 2.77) |
| Loukas, et al. <sup>9</sup> | 114/568 | 168/1,190 | 2.72 (2.10, 3.53) | 1.36 (1.01, 1.83) |
| Lozano, et al. <sup>38</sup> | 86/216 | 950/4,479 | 2.46 (1.85, 3.26) | 1.60 (1.31, 1.97)<br>** |
| Miech, et al. <sup>7</sup> | 4/13 | 14/213 | 6.32 (1.73, 23.10) | 6.58 (2.04, 57.88)<br>** |
| Morgenstern, et al. <sup>17</sup> | 93/312 | 175/1,867 | 4.11 (3.08, 5.48) | 2.50 (1.82, 3.54)<br>** |
| Primack, et al. <sup>20</sup> | 6/16 | 65/678 | 5.66 (1.99, 16.07) | 8.3 (1.2, 58.6) |
| Primack, et al. <sup>19</sup> | 6/16 | 81/889 | 6.06 (2.15, 17.10) | 6.82 (1.65, 28.25) |
| Spindle, et al. <sup>11</sup> | 45/153 | 230/2,163 | 3.50 (2.41, 5.09) | 3.37 (1.91, 5.94) |
| Treur, et al. <sup>13</sup> | 432/740 | 235/2,049 | 10.83 (8.87, 13.22) | 11.9 (3.36, 42.11) |
| Watkins, et al. <sup>12</sup> | 81/425 | 387/9,923 | 5.80 (4.46, 7.54) | 2.53 (1.80, 3.56) |
| Wills, et al. <sup>14</sup> | 42/215 | 50/926 | 4.25 (2.74, 6.61) | 2.87 (2.03, 4.05) |
| Overall | 1,451/4,787 | 4,340/52,727 | 4.59 (3.60, 5.85) | 2.92 (2.30, 3.71) |

\*Raw data were not available or insufficient information was provided to calculate an accurate unadjusted odds ratio.
\*\*Estimates were provided as risk ratios and converted to odds ratios.

**Figure 2.**
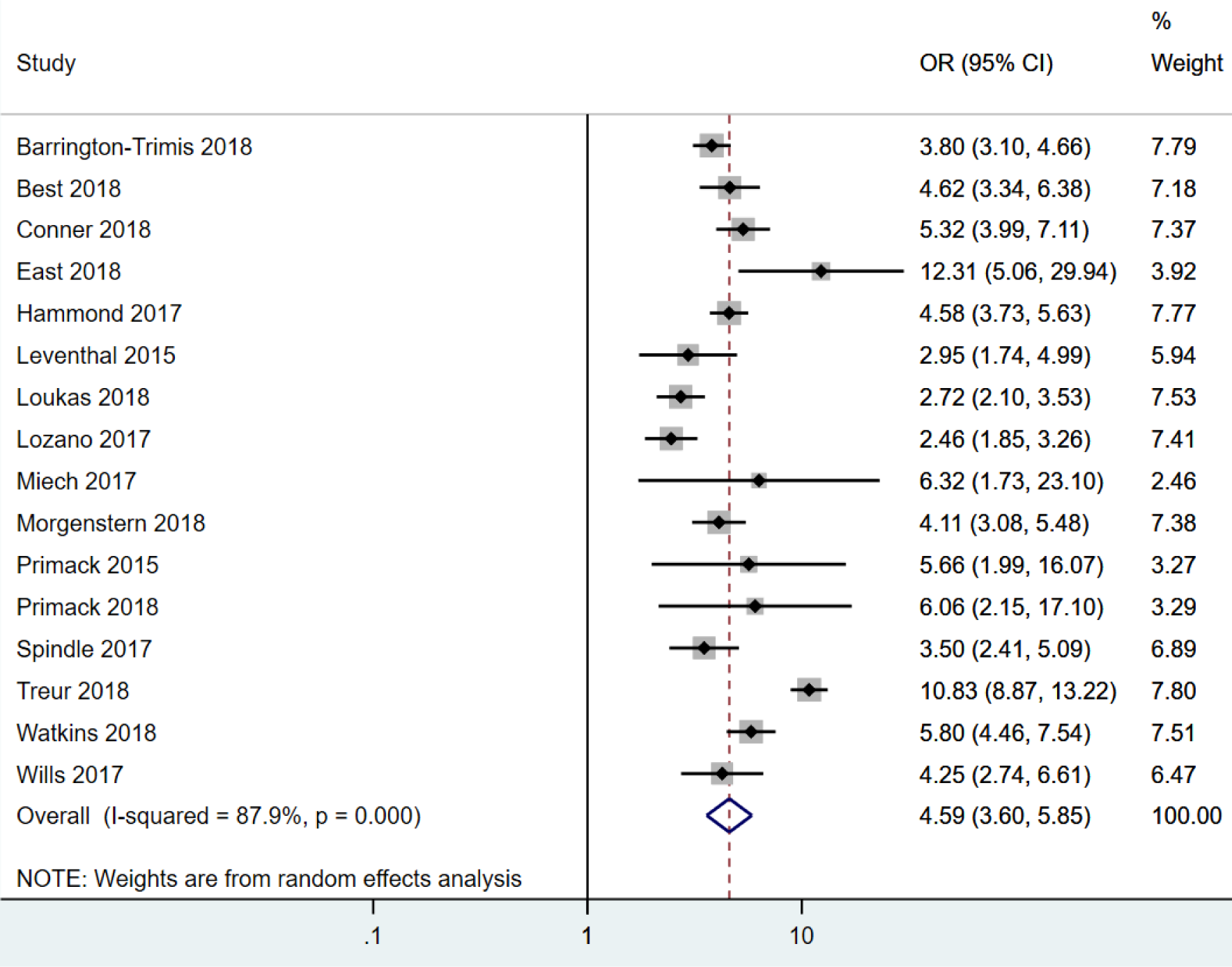
Forest plot for the unadjusted association between e-cigarette use and subsequent smoking.

**Figure 3.**
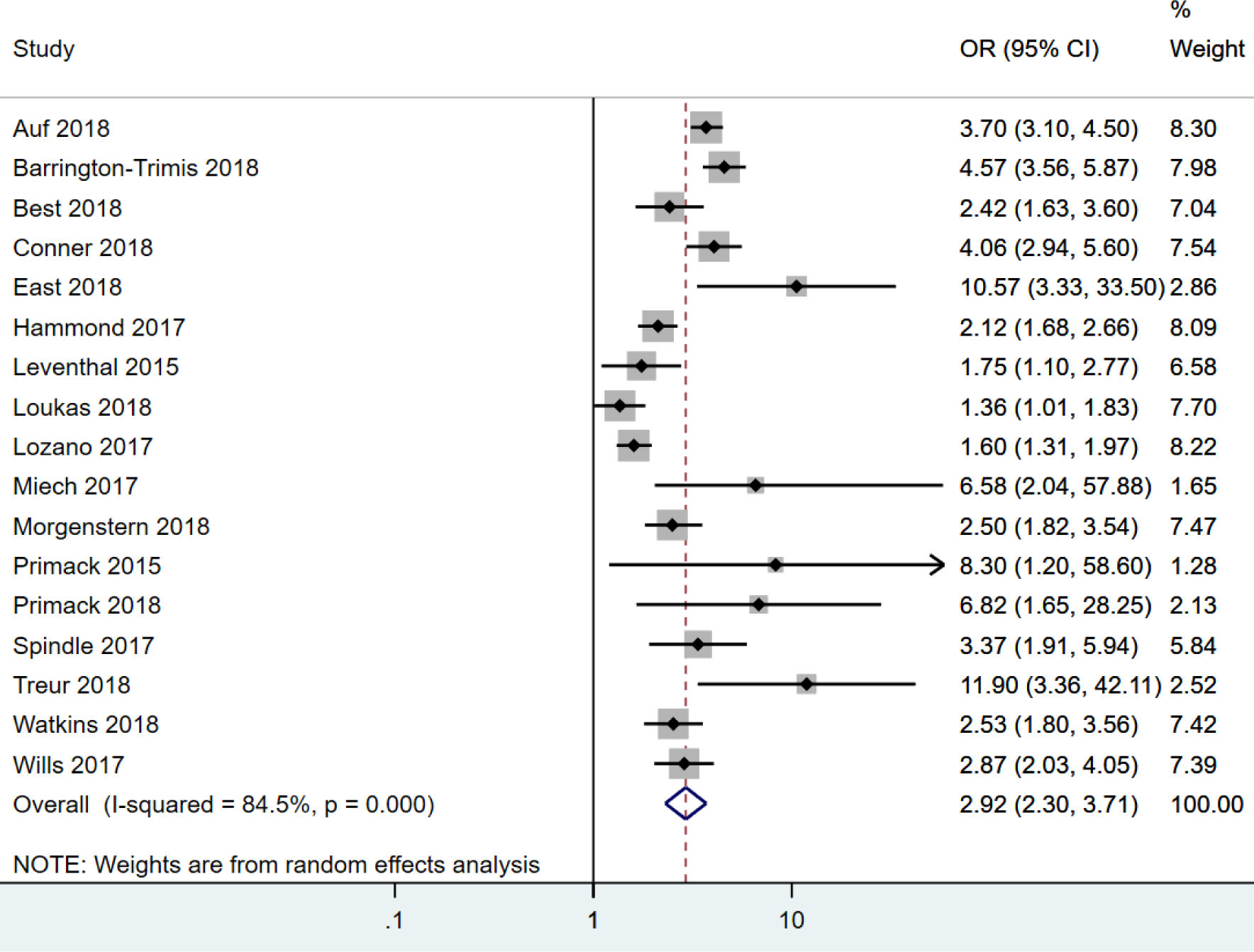
Forest plot for the adjusted association between e-cigarette use and subsequent smoking.

### Synthesis of Results

When pooled in a random effects meta-analysis, e-cigarette use in non-smoking young people was associated with a 4-and-a-half-fold increase in the odds of subsequent smoking (unadjusted; OR = 4.59, 95% CI 3.60 to 5.85). Pooling the adjusted estimates, the association was still strong but somewhat weaker (adjusted; OR = 2.92, 95% CI 2.30 to 3.71). Heterogeneity statistics indicated there was high heterogeneity in both the unadjusted (*I*^*2*^ = 88%) and adjusted (*I*^*2*^ = 85%) analyses.

Forest plots of the analyses sub-grouped by varying exposure and outcome levels can be found in the supplementary material (Figures S1-S4). Of the 16 studies included in the unadjusted meta-analyses, 13 provided results which explored ever e-cigarette use and ever smoking and three studies provided results for ever e-cigarette use and current smoking. Three studies explored past 30-day e-cigarette use and ever smoking. Further studies explored: past 30-day use of e-cigarettes and past 30-day smoking (1 study), frequency of e-cigarette use (1 study) and frequency of smoking (1 study). Pooled analyses were not possible for these subgroups.

### Ever e-cigarette use and ever smoking subgrouping

Pooled analyses of studies exploring ever vaping among never smokers and subsequent ever smoking resulted in a pooled unadjusted odds ratio of 4.17 (95% CI 3.53 to 6.29). Heterogeneity between included studies in this analysis was high (*I*^*2*^ = 90%). The results of the pooled adjusted analysis were similar with slightly lower odds (OR = 3.13, 95% CI 2.35 to 4.16). Heterogeneity between included studies was still high in the adjusted analysis (*I*^*2*^ = 84%).

### Ever e-cigarette use and current smoking subgrouping

Among never smokers, pooled unadjusted analyses indicated ever e-cigarette users had increased odds of subsequently becoming a current smoker (OR = 4.35, 95% CI 2.95 to 6.42) compared to never e-cigarette users. Heterogeneity estimates indicated low heterogeneity (*I*^*2*^ = 41%). Adjusted analyses showed similar but weakened results when pooled (OR = 2.12, 95% CI 1.72 to 2.84). Heterogeneity was indicated as low in the adjusted pooled analyses (*I*^*2*^ = 5%).

### Current e-cigarette use and ever smoking subgrouping

Past 30-day use of e-cigarettes among never smokers was associated with increased odds of ever subsequently smoking (OR = 5.64, 95% CI 3.75 to 8.50) in pooled unadjusted analysis. Heterogeneity estimates indicated low heterogeneity (*I*^*2*^ = 49%). The pooled adjusted analysis also indicated increased odds of ever subsequently smoking (OR = 2.33, 95% CI 1.84 to 2.96). Heterogeneity estimates also indicated low heterogeneity when the adjusted results were pooled (*I*^*2*^ = 5%).

### Stratification by age

When the unadjusted main analyses were stratified by age (including vs. excluding under 18 year olds) the pooled odds ratio among studies including those under 18 years was slightly higher (OR = 4.87, 95% CI 3.73 to 6.35) than the pooled odds ratio of studies excluding those under 18 (OR = 3.17, 95% CI 2.37 to 4.25). Heterogeneity estimates indicated that there was low heterogeneity between studies excluding under 18’s (*I*^*2*^ = 32%) but high heterogeneity between studies including under 18’s (*I*^*2*^ = 88%). Adjusted pooled analyses are reported in Supplementary Figures S5-S6.

### Stratification by study quality/risk of bias

Due to limited variation in quality rating of studies using the NOS, fair and poor quality studies were pooled and compared to good quality studies. The pooled unadjusted odds ratio for studies rated as good quality (n = 13 studies; OR = 4.29, 95% CI 3.67 to 5.01) was lower than the pooled odds ratios for fair/poor quality studies (n = 3 studies; OR = 5.41, 95% CI 1.67 to 17.51). Heterogeneity measures indicated that high quality studies were less heterogeneous than fair/poor quality studies (*I*^*2*^ = 60% and *I*^*2*^ = 97% respectively). Adjusted pooled analyses are shown in Supplementary Figures S7-S8.

### Stratification by support for the gateway hypothesis

During the review process it became apparent that many studies did not draw clear conclusions regarding the gateway hypothesis or made balanced conclusions. This made it difficult to categorise studies and as such we were unable to stratify based on this criterion.

### Stratification by location of study

There were only two countries in which more than one included study was conducted; 10 studies took place in the US and three took place in the UK. The pooled estimate for unadjusted odds of studies conducted in the US was 3.95 (95% CI 3.17 to 4.92) and in the UK was 5.55 (95% CI 3.94 to 7.82). Heterogeneity for studies conducted was high in the US (*I*^*2*^ = 93%) and moderate in the UK (*I*^*2*^ = 52%). The other studies were located in the Netherlands, Germany, Canada and Mexico. Pooling these results, the odds of subsequent smoking was 4.75 (95% CI 2.54 to 8.89) with high heterogeneity between studies (*I*^*2*^ = 96%). Adjusted pooled analyses are shown in Supplementary Figures S9-S11.

### Risk of Bias Across Studies

The risk of bias across studies is shown in a funnel plot in Figure 4. The figure is somewhat asymmetrical and some points (25%) lie above the superimposed funnel limits (95% confidence region) suggesting that there may be some publication bias and indicating there may be heterogeneity (as supported by the *I*^2^ statistics) or selection bias across the included studies.^21^

**Figure 4.**
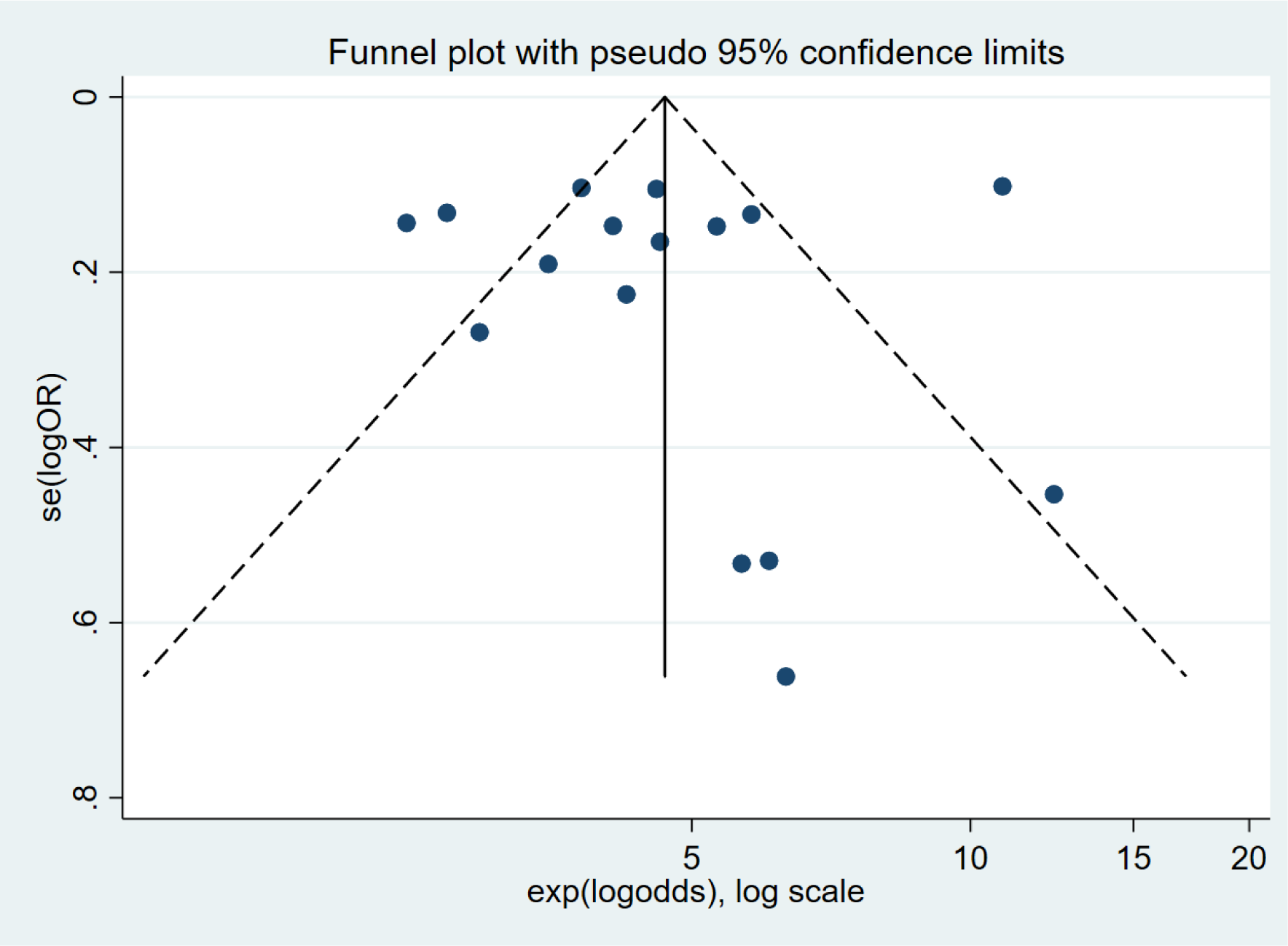
Funnel plot to assess risk of bias across studies.

## Discussion

Our results indicated that self-reported non-smokers who have used e-cigarettes have 4-and-a-half-fold higher odds of subsequently reporting being smokers than those who have not. The pooled adjusted estimate indicated a weaker, although still strong, association with nearly 3-fold increased odds of later smoking. Sub-grouping and stratification revealed some differences between groups, but all findings indicated a strong positive association. The main findings were consistent with the findings of Soneji and colleagues,^5^ whereby there was a strong positive association between e-cigarette use among non-smokers and subsequent smoking and a high degree of heterogeneity between studies. Similar to Soneji and colleagues,^5^ stratification by age revealed slightly lower pooled estimates for the odds of smoking in studies which excluded under 18-year olds compared to studies including them. Stratification by location indicated stronger associations in the UK compared to the US and other countries. We identified an additional 11 studies which were not included in the Soneji meta-analysis. One study which was included in the Soneji and colleagues meta-analysis^5^ was not included in this meta-analysis; it was substituted for more recent evidence using an overlapping data set.^22^ In addition, one conference abstract was substituted for the published article.^23^

Three of the four pre-selected Bradford-Hill criteria (strength of association. temporality and specificity) were commonly rated as having been met in the included studies. Despite the high heterogeneity statistics, the included study estimates are consistently in the same direction and there are plausible causal pathways (e.g., nicotine addiction, similar hand-to-mouth actions for both behaviours). This suggests the results provide some support for a causal relationship between e-cigarette use and later smoking. This is in line with the theory that e-cigarettes act as a gateway to smoking.^3 24^ Commonly in the literature, the claim is that the gateway effect is attributable to nicotine addiction.^24^ E-cigarettes have historically not delivered nicotine as effectively as cigarettes,^25^ so that e-cigarettes may not be adequate to satisfy users who become more heavily addicted to nicotine.^26^ In contrast, the common liability theory proposes that people who use multiple drugs (or in this case different delivery methods of the same drug) share the same predisposing factors.^27^ When considering a causal relationship between e-cigarette use and later smoking (e.g., the gateway hypothesis), these shared factors/confounders (i.e., the common liability theory) should be taken into account. Due to a lack of consideration of such factors in the included studies, we have some reservations about inferring a causal relationship. We discuss our reservations below with respect to Bradford-Hill criteria for causality, highlighting the need for additional research which may help to strengthen the evidence for a causal relationship.

The majority of studies included in the meta-analysis satisfied the temporality criterion. All studies except one were longitudinal and measured exposure prior to smoking and smoking at follow up; therefore, the observed association between earlier e-cigarette use and later smoking is unlikely to be due to reverse causality (i.e., smoking leading to e-cigarette use). However, smoking status is sometimes misreported by young people,^28^ meaning that some self-reported non-smokers at baseline may in fact have smoked previously. If ever smokers misreport their smoking history at baseline, but they accurately report that they are ever smokers at follow up, the association could be biased away from the null. Self-reports were validated in one study ^29^ using breath carbon monoxide levels; however, with the short half-life of breath carbon monoxide (4-6 hours) this measure is not suitable to validate self-reports of ever smoking. In future research, the use of biomarkers which can identify long term exposure could help researchers to objectively confirm self-reported smoking status at baseline. Differential DNA methylation signals have been observed among smokers compared to non-smokers ^30 31^and it is plausible that different signals would be observed among e-cigarette users. Eventually, researchers may be able to use these methylation signatures to exclude people who misreport their behaviour. Consequently, researchers could more confidently dismiss reverse causation as an explanation for the association.

It is also worth noting that our measure of specificity was relatively liberal. We rated studies as specific if they adjusted for more than basic demographic factors. In a recent cross-sectional study, accounting for shared risk factors fully explained the relationship between e-cigarette use and current smoking, demonstrating the importance of adjusting for potential confounders such as alcohol and drug use, peer smoking and risk taking behaviour.^32^ In addition, some potential confounders, like impulsivity, are difficult to fully capture via self-report and are often assessed relatively crudely. Had we considered only studies adjusting for behavioural risk factors (e.g., alcohol and marijuana use) to meet this criterion, only six studies would have been rated as specific. Adjusting for these factors considerably reduced the odds ratio and/or widened the confidence interval compared to the unadjusted analysis. Since statistical adjustment can never fully remove the risk of confounding, other approaches to exploring the potential for a common liability (e.g., to risk taking) explaining the observed association between e-cigarette use and smoking are warranted. One way of exploring specificity of this relationship more thoroughly (which none of the included studies did) would be the use of negative control outcomes (i.e. outcomes which have similar confounding structures to smoking but for which there is no biologically plausible mechanism for e-cigarette use being a causal factor). For example, using an e-cigarette is unlikely to cause other risky behaviours such as the number of sexual partners a person has; if similar associations are seen between e-cigarette use and both smoking and number of sexual partners, it would indicate that the link may be caused by common underlying factors. Furthermore, exploring the genetic aetiology of e-cigarette use may help in understanding whether e-cigarette use and smoking share a common liability. If e-cigarette use has a shared genetic aetiology with negative control outcomes for which there are no plausible pathways through which e-cigarette use would be a causal factor (e.g., gambling) this would suggest that the association is due to the two behaviours sharing a common genetic liability. The triangulation of evidence obtained using different methods will be critical here.^33^

To meet the final pre-selected Bradford-Hill criterion, dose-response, increased e-cigarette use should lead to greater risk of later smoking. Despite the most likely causal pathway from e-cigarette use to later smoking being via nicotine addiction,^3^ only one of the included studies measured and took into account the nicotine content of the e-cigarettes used. This study indicated that both use of nicotine containing e-cigarettes and (to a lesser extent) non-nicotine containing e-cigarettes are strongly associated with later smoking.^13^ This suggests that nicotine exposure may be one factor in the association between e-cigarette use and later smoking, but not the sole mechanism.

Unfortunately, the study reported analyses based on nicotine vs non-nicotine vaping in which these two groups were not mutually exclusive (i.e., individuals would be in both analysis groups if they tried both nicotine containing and nicotine free e-cigarettes).

Thus, it is unclear whether there is an association between e-cigarette use among non-smokers and later smoking when users have not been exposed to nicotine. To determine whether there is a nicotine dose-response involved in the association, we would also need to observe the frequency of e-cigarette use prior to smoking. The one study that looked at frequency of use (at 4 levels) indicated that there may be a dose-response to nicotine when comparing use just once/twice to use weekly/daily.^14^ However, the odds of later smoking did not increase linearly with each increased level of frequency of use. However, as nicotine is metabolised differently on an individual basis, direct measures of nicotine rather than frequency of use may be necessary to determine whether there is a dose-response. Although nicotine may play a causal role in the relationship between e-cigarette use and smoking, without further study of any dose-response relationship (including the study of nicotine content and frequency of e-cigarette use), we cannot confidently infer causality according to Bradford-Hill criteria.

Our heterogeneity statistics of 88% (unadjusted analyses) and 85% (adjusted analyses) indicate that our effect estimates should be interpreted with caution. Although there is no commonly agreed upon strict threshold for heterogeneity, as a rough guide the Cochrane handbook suggests that *I*^2^ statistics of between 75% and 100% represent considerable heterogeneity.^8^ Age contributed to the observed heterogeneity in our meta-analysis – the association was stronger in studies including under 18 year olds than studies excluding them. In adolescence, risk-taking is common ^34^ and decision making for health-risk behaviours is influenced by peers, societal influences and parental monitoring, but these factors are less influential to adults.^35^ Such factors are likely to be confounders of the association between e-cigarette use and later smoking, particularly in studies of under 18 year olds and should be included as covariates where possible.

The results stratified by location also suggested there may be societal influences on the association; the association was stronger among studies based in the UK than those based in the US. This suggests that country-specific societal factors such as, legislation, taxation, social norms, and public opinion may be confounding this association such that study results may not be generalisable to other countries.

In conclusion, there is a strong consistent association in observational studies between e-cigarette use among non-smokers and later smoking. However, findings from published studies do not provide clear evidence that this is explained by a gateway effect rather than shared common causes of both e-cigarette use and smoking. This emphasises the need for a scientific forum to discuss the evidence to date and directions for future research. Future research should consider including relevant potential confounders, such as better measures of impulsivity and other measures of propensity to risk taking, and objective measures of smoking status in order to better explore the potential role of e-cigarettes as a gateway to smoking. Studies that explore the genetic underpinnings of these behaviours and use negative control outcomes may also help improve our understanding of the association between e-cigarette use and later smoking. A scoping review, including qualitative evidence, could provide a clearer understanding of the why e-cigarette use is associated with later smoking. Importantly, any recommendations regarding e-cigarette regulations to limit the burden of future smoking must consider the potential beneficial impact of e-cigarette use on smoking cessation.^36^

## Data Availability

All data has been made available in the supplementary material.

## Contributors

JNK, AET and MRM conceived and designed the study. JNK, SFS and SP acquired and analysed the data, and JNK, AET and MRM interpreted the data. JNK wrote the initial draft of the manuscript and all the authors were involved in preparing this manuscript and contributed to the revision of the manuscript. JNK, AET and MRM are guarantors. The corresponding author attests that all listed authors meet authorship criteria and that no others meeting the criteria were omitted.

## Ethical approval

Ethical approval was not obtained for this study as approval was obtained by the original authors of the included studies.

## Data sharing

The datasets used in this analysis are available on request.

